# An Early Adverse Drug Event Detection Approach with False Discovery Rate Control

**DOI:** 10.1101/2023.05.31.23290792

**Authors:** Yi Shi, Xueqiao Peng, Ruoqi Liu, Anna Sun, Yuedi Yang, Ping Zhang, Pengyue Zhang

## Abstract

Adverse drug event (ADE) is a significant challenge in clinical practice. Many ADEs have not been identified timely after the approval of the corresponding drugs. Despite the use of drug similarity network demonstrates early success on improving ADE detection, false discovery rate (FDR) control remains unclear in its application. Additionally, performance of early ADE detection has not been explicitly investigated under the time-to-event framework. In this manuscript, we propose to use the drug similarity based posterior probability of null hypothesis for early ADE detection. The proposed approach is also able to control FDR for monitoring a large number of ADEs of multiple drugs. The proposed approach outperforms existing approaches on mining labeled ADEs in the US FDA’s Adverse Event Reporting System (FAERS) data, especially in the first few years after the drug initial reporting time. Additionally, the proposed approach is able to identify more labeled ADEs and has significantly lower time to ADE detection. In simulation study, the proposed approach demonstrates proper FDR control, as well as has better true positive rate and an excellent true negative rate. In our exemplified FAERS analysis, the proposed approach detects new ADE signals and identifies ADE signals in a timelier fashion than existing approach. In conclusion, the proposed approach is able to both reduce the time and improve the FDR control for ADE detection.

## 1. Introduction

Adverse drug event (ADE) is defined as any drug exposure related harmful patient experience.^1^ ADE is considered as a significant challenge in current clinical practice.^2-5^ For instance, it is estimated that ADEs cause as many as 4.5 million ambulatory encounters, 1.3 million emergency department visits, 350,000 hospitalizations, and 106,000 deaths, in the United States (US) each year.^6-8^ Many ADEs have been identified years after the corresponding drugs’ approval dates. For instance, in the US, the periods from approval to withdrawal due to safety concerns were 3.4 years for valdecoxib, 4.7 years for tegaserod, and 5.4 years for efalizumab.^9^ These facts necessitate post-marketing pharmacovigilance for drug safety.^10^

To promote pharmacovigilance, many regulatory agencies maintain spontaneous reporting systems (SRSs) to collect ADE reports. For instance, the US Food and Drug Administration’s (FDA’s) Adverse Event Reporting System (FAERS) is a large-scale SRS database including over 24 million ADE reports.^11^ Signal detection algorithms (SDAs) have been developed to identify ADE signals from SRS databases. Frequentist SDAs include proportional reporting ration (PRR),^12^ reporting odds ratio (ROR),^13^ and likelihood ratio test (LRT).^14^ The gamma Poisson Shrinker (GPS) and the Bayesian confidence propagation neural network (BCPNN) are empirical Bayes SDA and Bayesian SDA, respectively.^15,16^ For a particular drug-ADE pair, SDAs summarize all ADE reports into a 2-by-2 contingency table according to the presence of the drug (yes/no) and the ADE (yes/no). Subsequently, an ADE signal is measured by variants of the observed frequency to expected frequency ratio, where the expected frequency is computed under the assumption of no drug-ADE association. Comparing to frequentist SDAs, GPS and BCPNN are able to penalize false ADE signals generated by drugs with a low report frequency; and they have been routinely used by the FDA and the World Health Organization (WHO).^17^ Additionally, Ahmed *et al*. used posterior probability of null hypothesis under the GPS model to control false discovery rate (FDR) for high-throughput ADE mining (e.g. simultaneously mining over a million of drug-ADE pairs).^18^ Tatonetti *et al*. used multiple logistic regression to generate confounder-adjusted ADE signals.^19^ The ADE signals identified by SDAs had their own validations, and many promising discoveries were successfully validated.^20,21^ Despite the successes of SDAs, signal detection shall be expand to identify ADE signals both in a timelier fashion and at a low FDR.^22^ Recent studies utilized drug similarity network to enhance existing SDAs.^23-25^ For instance, Liu and Zhang used drug similarity network to enhance signals generated from PRR, ROR, GPS and BCPNN.^24^ Ji *et al*. enhanced signals from BCPNN by leveraging drug similarity network derived prior ADE risk distribution.^23^ While enhanced ADE signals demonstrated improved performance, FDR control remains unclear on prioritizing these signals. Additionally, the performance of early ADE detection has not been explicitly investigated under a time-to-event setting (e.g., time from first report to time of ADE detection).

In this manuscript, we propose an early ADE detection approach. Specifically, under the proposed approach, prior ADE risk distribution of a new drug can be derived from ADE risks of existing drugs with ample ADE reports and similar chemical structures to the new drug. Subsequently, the drug similarity based posterior probability of null hypothesis for testing the new drug’s ADE risk can be computed from the new drug’s observed data and prior ADE distribution. The drug similarity based posterior probability is able to both facilitate early ADE detection and provide much desired FDR control for monitoring a large number of ADEs of multiple new drugs. The rest of the manuscript is organized as following. Section 2 describes the datasets and approaches. Section 3 includes performance evaluation analysis, simulation study and an exemplified FAERS analysis. Section 4 presents our conclusion and discussion.

## 2 Method

### 2.1 Data Preparation

#### 2.1.1 Drug Similarity Network

We used 881 chemical substructures defined in Pubchem to construct the drug similarity network.^26^ We assigned an 881-component vector to each drug, where a component equals to 1 or 0 corresponding to the presence or absence the substructure. Let 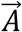 and 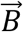 denote vectors of two drugs, respectively. We used the Jaccard Score to calculate the chemical similarity of the drug-drug pair:

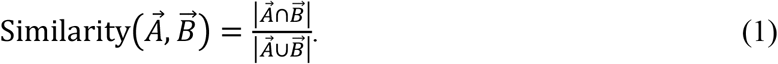

We computed the similarity scores for all drug-drug pairs. The 75% percentile of pairwise drug similarities was 0.44 for all pairs. The 75% percentile of pairwise drug similarities was 0.62 for pairs sharing a level 3 class of the Anatomical Therapeutic Chemical (ATC) Classification (e.g., drugs acting on similar systems).^27^

#### 2.1.2 FAERS Data

We followed the protocol described in Banda et al. to process the FAERS data (years: 2004-2018).^28^ Specifically, we removed duplicate records, normalized drug names to ingredient names according to RxNorm and DrugBank ID,^29,30^ and mapped ADE names to Medical Dictionary for Regulatory Activities (MedDRA) terms.^31^ We excluded ADE reports with more than ten ingredient names. Our final dataset included 8.68 million reports, 2,453 ingredient names with chemical similarity data, and 10,711 MedDRA PT/LLT terms with report frequencies ≥100.

#### 2.1.3 Drug Label Data

We used two ADE datasets curated from drug labels for performance evaluation (e.g., Demner-Fushman et al. and SIDER).^32,33^ MedDRA terms were available in both Demner-Fushman et al. and SIDER.^32,33^ For both ADE datasets, we included drugs had: 1) reporting frequency =0 on 2004Q1 (e.g., “new drugs” on 2004Q1); and 2) ≥20 drugs with similarity scores ≥0.6 and reporting frequencies ≥2,000 when the “new drugs” were firstly reported. The rationale of using “new drugs” is to facilitate the evaluation of early ADE detection. We identified 46 new drugs and 2,711 labeled drug-ADE pairs from Demner-Fushman et al.^33^ We identified 53 new drugs and 4,582 labeled drug-ADE pairs from SIDER.^32^

### 2.2 Adverse Drug Event Detection

#### 2.2.1 Notations

For a new drug and an ADE, let *N* and *X* be the number of reports involving the drug and the drug-ADE pair, respectively. Let *θ* be the ADE reporting rate of the drug. Let 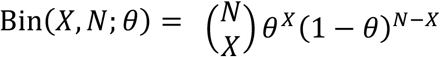 denote the probability mass function (PMF) of binomial distribution (*X* = 0, …, *N* and 0 < *θ* < 1). Let 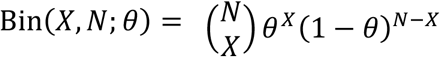 denote the probability density function (PDF) of the beta distribution (0 < *θ* < 1, *α* > 0 and *β* > 0). Let 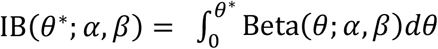 denote the incomplete beta distribution. Additionally, let subscript *i* indicate the *i*th drug (*i* = 1, …, *I*), let subscript *j* indicate the *j*th ADE (*j* = 1, …, *J*), and let subscript (*k*) indicate the *k*th time point (*k* *≥* 1).

#### 2.2.2 Methods to Detect ADE

The following test can be used to detect the *j*th ADE of the *i*th new drug:

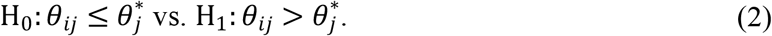

In equation (2), 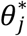 is a predetermined value. For instance, 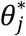 can be selected as the marginal reporting rate of the *j*th ADE. We assume *X*_*ij*_∼Bin(*X*_*ij*_, *N*_*ij*_; *θ*_*ij*_) and *θ*_*ij*_∼Beta (*θ*_*ij*_; *α*_*ij*_, *β*_*ij*_) under the Bayesian framework. In FAERS analysis, *α*_*ij*_ and *β*_*ij*_ in Beta(*θ*_*ii*_; *α*_*ii*_, *β*_*ii*_) can be estimated from ADE reports of existing drugs that have high chemical similarities with the new drug. The posterior probability of null hypothesis (2) given *X*_*ij*_ and *N*_*ij*_ can be expressed as

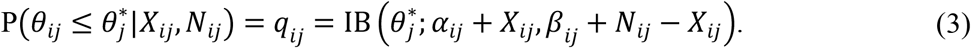

Let *c* denote the threshold of *q*_*ij*_. The decision rule for using the posterior probability of null hypothesis to detect ADE is

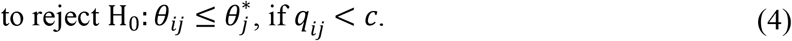

Naturally, *q*_*ij*_ measures the false discovery portion (FDP) if *q*_*ij*_ < *c*.^18^

The decision rule (4) can be used to control the FDR for high-throughput ADE mining. As described in Ahmed et al.,^18^ The FDR of mining *I* × *J* drug-ADE pairs given a selected threshold *c* can be expressed as:

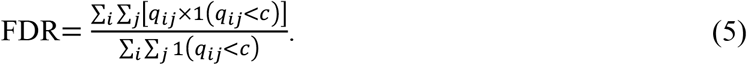

Thus, *c* controls FDR naturally, as FDR < *c* for any given *c*.

The decision rule (4) can be generalized to sequentially monitoring a large number of ADEs of multiple new drugs. A fixed threshold *c* of the posterior probabilities (3) can be selected for the sequential monitoring approach.^34^ In FAERS analysis, ADE can be monitored at each fiscal quarter. The decision rule for the sequential monitoring approach is

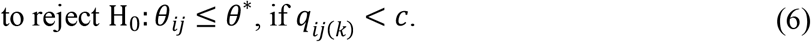

As posterior probabilities are not affected by the data collection process, the decision rule (6) is able to control FDR at *c* for sequentially monitoring all potential ADEs of multiple new drugs.

### 2.3 Methods Used in Performance Evaluation Analysis

FDR control for high-throughput ADE mining was firstly defined in Ahmed et al. under the GPS model.^18^ For the *i*th drug and *j*th ADE, let *X*_*ij*_ be its observed frequency and *E*_*ij*_ be its expected frequency by assuming no drug-ADE association (e.g., *E*_*ij*_ = *X*_*i*+_*X*_+*j*_/*X*_++_, where *X*_*i*+_, *X*_+*j*_ and *X*_++_ are the marginal drug reporting frequency, the marginal ADE reporting frequency and the overall reporting frequency, respectively). Under the GPS model, the underlining relative risk (i.e., *λ*_*ij*_) of the drug-ADE pair follows a mixture of gamma distribution given *X*_*ij*_ and *E*_*ij*_:

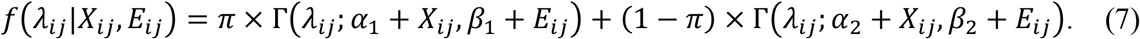

From (7), the posterior probability of null hypothesis under the GPS model can be expressed as:

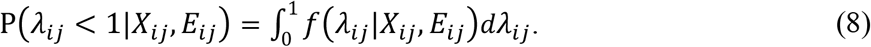

Under the BCPNN model, the report frequencies of the *i* th drug, the *j* th ADE and the corresponding drug-ADE pair are assumed to follow binomial distributions. The probabilities in the binomial distributions are further assumed to follow uniform or beta distributions:

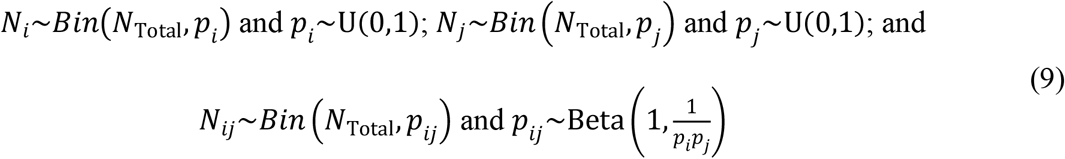

Under the BCPNN model, drug-ADE pair can be ranked by the expectation of the information component (IC).^15^

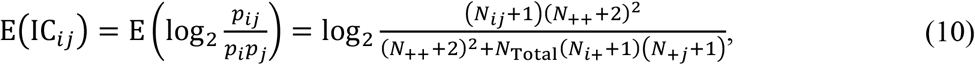

where *X*_*i*+_, *X*_+*j*_ and *X*_++_ are the marginal drug reporting frequency, the marginal ADE reporting frequency, and the overall reporting frequency, respectively. Posterior probability and/or FDP estimation is not analytically available for the BCPNN model.^18^

## 3 Results

### 3.1 Performance Evaluation Analysis Using Drug Label Data

We compared the proposed drug similarity based posterior probability of null hypothesis to: 1) the posterior probability of null hypothesis under the GPS model; and 2) the expectation of the information component (IC) under the BCPNN model. We analyzed the FAERS data with respect to 46 and 53 “new drugs” in Demner-Fushman et al. and SIDER (please see section 2.1.3),^32,33^ respectively. Under the proposed approach, each “new drug’s prior ADE risk distribution was derived from the top-20 similar drugs that had report frequency ⩾2000 at the time that the corresponding “new drug” was firstly reported (i.e., first report year/quarter). We monitored all ADEs with marginal report frequencies ⩾100 in the FAERS data for the “new drugs”. For each drug-ADE pair, we computed: 1) the proposed drug similarity based posterior probability of null hypothesis (e.g., PP_sim); and 2) the posterior probability of null hypothesis under the GPS model (e.g., PP_GPS); and the expectation of IC under the BCPNN model (e.g., BCPNN).

First, we compared the area under the receiver operating characteristic curve (AUC) values for all approaches. Specifically, we used labeled drug-ADE pairs as condition positives and unlabeled drug-ADE pairs as condition negatives. Subsequently, we computed AUC values for PP_sim, PP_GPS, and BCPNN at each quarter after the first reporting time for each of the “new drugs”. Figure 1A and Figure 1B illustrate the AUC values up to 7 years after the first reporting time. Specifically, in Figure 1A and Figure 1B, the lines represent median AUC values for all “new drugs” at each year/quarter, and the boxes represents the average of the 5%, 25%, 50%, 75%, and 95% quantiles of the AUC values for all “new drugs” at each year. Using ADEs in Demner-Fushman et al.;^33^ we observed the proposed PP_sim had higher AUC values compared with PP_GPS, and BCPNN within 2 years after the new drugs were firstly reported (median AUC values at year 2: PP_sim =0.66, PP_GPS =0.61, and BCPNN =0.60); while all approaches had comparable AUC values at 4 years after the new drugs were firstly reported (median AUC values at yea 4r: PP_sim =0.72, PP_GPS =0.69, and BCPNN =0.71). Using ADEs in SIDER,^32^ we observed the proposed PP_sim had higher median AUC values compared with PP_GPS, and BCPNN within 2 years after the new drugs were firstly reported (median AUC values at year 2: PP_sim =0.62, PP_GPS =0.55, and BCPNN =0.54); and at 4 years after the new drugs were firstly reported (AUC values at year 4: PP_sim =0.64, PP_GPS =0.61, and BCPNN =0.61).

**Figure 1.**
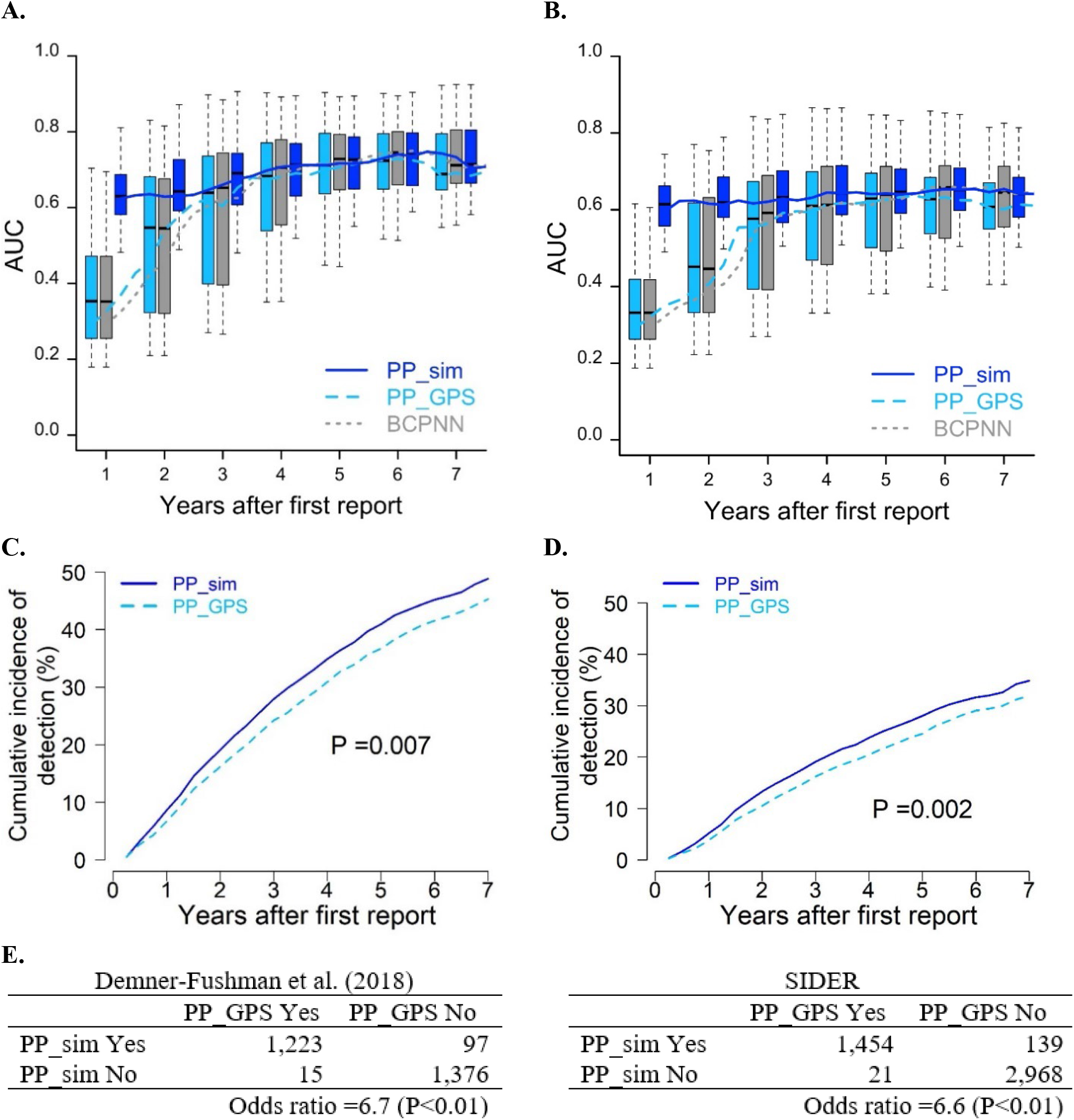
Performance evaluation analysis using FAERS data and label adverse drug events (ADEs); **A**. Area under the receiver operating characteristic curve (AUC) values after first drug reporting time for ADEs in Demner-Fushman et al. (2018); **B**. AUC values after first drug reporting time for ADEs in SIDER; **C**. Cumulative incidence from first reporting time to detection for ADEs in Demner-Fushman et al. (2018); **D**. Cumulative incidence from first reporting time to detection for ADEs in SIDER; **E**. Comparison of detected ADEs in Demner-Fushman et al. (2018) and SIDER.

Second, we examined the time to detection of labeled ADEs under the time-to-event setting. As FDR control was unavailable under the BCPNN model, we examined PP_sim and PP_GPS by using 0.05 as the threshold of the posterior probabilities, which was equivalent to control FDR at 0.05. Figure 1C and Figure 1D illustrate the cumulative incidence of detection. We observed the proposed PP_sim was able to detect ADE earlier than PP_GPS (P-values ⩽0.002 under log-rank test; [Figure 1C and Figure 1D]). Additionally, PP_sim was able to detect more labeled ADEs than PP_GPS (Figure 1E). As Figure 1E shown, the proposed PP_sim had odds ratios (ORs) ⩾6.5 (Ps <0.01 under the McNemar’s test) comparing to PP_GPS for identifying labeled ADEs. Last, we investigated the prior ADE risk distributions. The median of the minimal similarity value between a new drug and its similar drugs was 0.64 (interquartile range [IQR]: 0.63 - 0.66). We observed the labeled ADEs had significant smaller prior probabilities of the null hypothesis compared with the unlabeled ADEs (P-values <0.001 under the Mann–Whitney *U* test [Figure S1]).

### 3.2 Simulation Study

We conducted simulation study to investigate the performance metrics of the proposed PP_sim and the PP_GPS, as both approaches focused on FDR control for mining a large number of ADEs. We simulated ADE report data of a new drug according to the characters of the FAERS data. Specifically, we assumed the “new drug” was firstly reported on 2009Q1. We assumed the new drug had high similarities with 23 drugs (tolterodine, diphenhydramine, duloxetine, paroxetine, fluoxetine, venlafaxine, levothyroxine, amiodarone, verapamil, gemfibrozil, tramadol, propranolol, atenolol, bisoprolol, metoprolol, salbutamol, salmeterol, citalopram, escitalopram, tamoxifen, tamsulosin, ramelteon, and lopinavir). All of these 23 selected drugs had report frequencies ⩾ 2,000 on 2009Q1. The minimal pairwise similarity between these 23 drugs was 0.65.

The simulation procedure included the following steps. First, we derived the new drug’s prior ADE reporting rate distributions for all ADEs by using the ADE reporting data of the aforementioned 23 drugs on 2009Q1. We simulated the new drug’s underlying ADE reporting rates from the prior distributions. We compared the underlying ADE reporting rates of the new drug to the corresponding marginal ADE reporting rates in FAERS. We defined true positives (i.e., underlying rate > marginal rate) and true negatives (i.e., underlying rate ⩽ marginal rate). Second, we assessed the quarterly reporting rates (e.g. accumulation rate of reports) after initial reporting for all drugs that were firstly reported on or after 2009Q1. We computed the median quarterly reporting frequencies for those drugs. We assumed the report frequencies of the simulated “new drug” to follow Poisson distribution with mean equaled to the aforementioned median accumulation rates in FAERS. We assumed the ADE frequencies to follow binomial distributions with the underlying ADE reporting rates of the simulated “new drug”. We simulated ADE reports of the new drugs up to 20 quarters (e.g., 5 years) after 2009Q1. We conducted 100,000 simulations. In each simulation, we computed PP_sim and PP_GPS. Further, we computed the empirical FDR, empirical true positive rate (TPR), empirical true negative rate (TNR) Based on 100,000 simulations.

Figure 2A shows the empirical FDRs. We observed both PP_sim and PP_GPS were able to control the FDR at desired level. Both PP_sim and PP_GPS had FDRs closed to 0 immediately after first reporting time. The empirical FDRs were peaked at year 1 after first reporting time (empirical FDRs at year 1: PP_sim =0.015 and PP_GPS =0.011). PP_GPS had conservative empirical FDR values than PP_sim.

**Figure 2.**
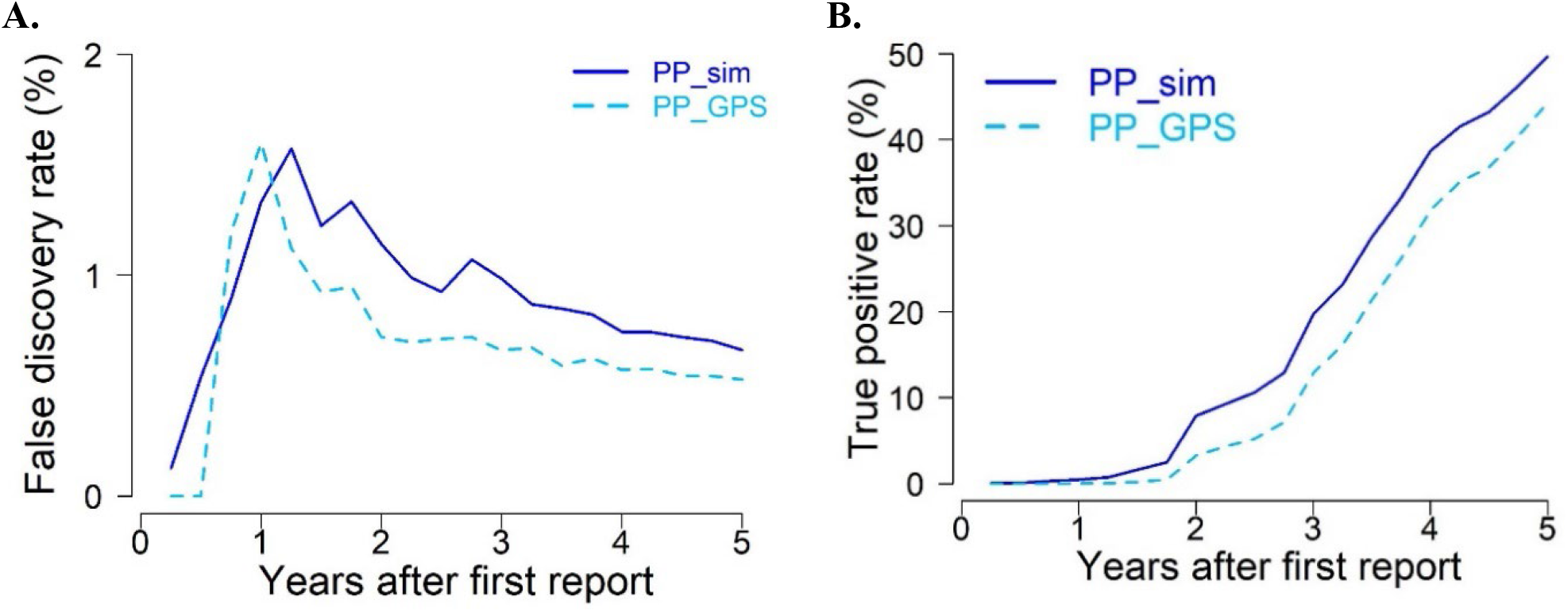
Simulation results; **A**. False discovery rate (FDR); **B**. True positive rate (TPR).

Figure 2B shows the empirical TPRs. Both approaches had empirical TPRs increase as time increase, while PP_sim had better empirical TPRs than PP_GPS (empirical TPRs at year 5: PP_sim =0.49 and PP_GPS =0.44). Both approaches had excellent empirical TNRs (empirical TNRs >0.99 in years 1 - 5 [Figure S2]).

### 3.3 FAERS Data Analysis

We analyzed 205 “new drugs” that were firstly reported on or after 2012Q1. We computed PP_sim and PP_GPS for 1.33 million drug-ADE pairs. By controlling FDR at 0.05, PP_sim and PP_GPS identified 10,650 ADE signals and 7,298 ADE signals, respectively. Of the 7,298 ADE signals identified by PP_GPS, 7,240 were also identified by PP_sim. In another word, 99.2% of the ADE signals identified by PP_GPS were also identified by PP_sim. Alternatively, PP_sim uniquely identified 3,410 ADE signals that had not been identified by PP_GPS.

Figure 3A shows the percentages of the ADE signals of PP_sim that were also identified by PP_GPS. The frequencies were stratified by drug reporting frequency and the marginal ADE reporting rate. As Figure 3A shown, PP_sim uniquely identified more ADEs signals with lower drug reporting frequencies and/or a smaller marginal ADE reporting rates. Further, for the 7,240 ADE signals identified by both approaches, we observed PP_sim had significant shorter time to detection than PP_GPS (Figure 3B).

**Figure 3.**
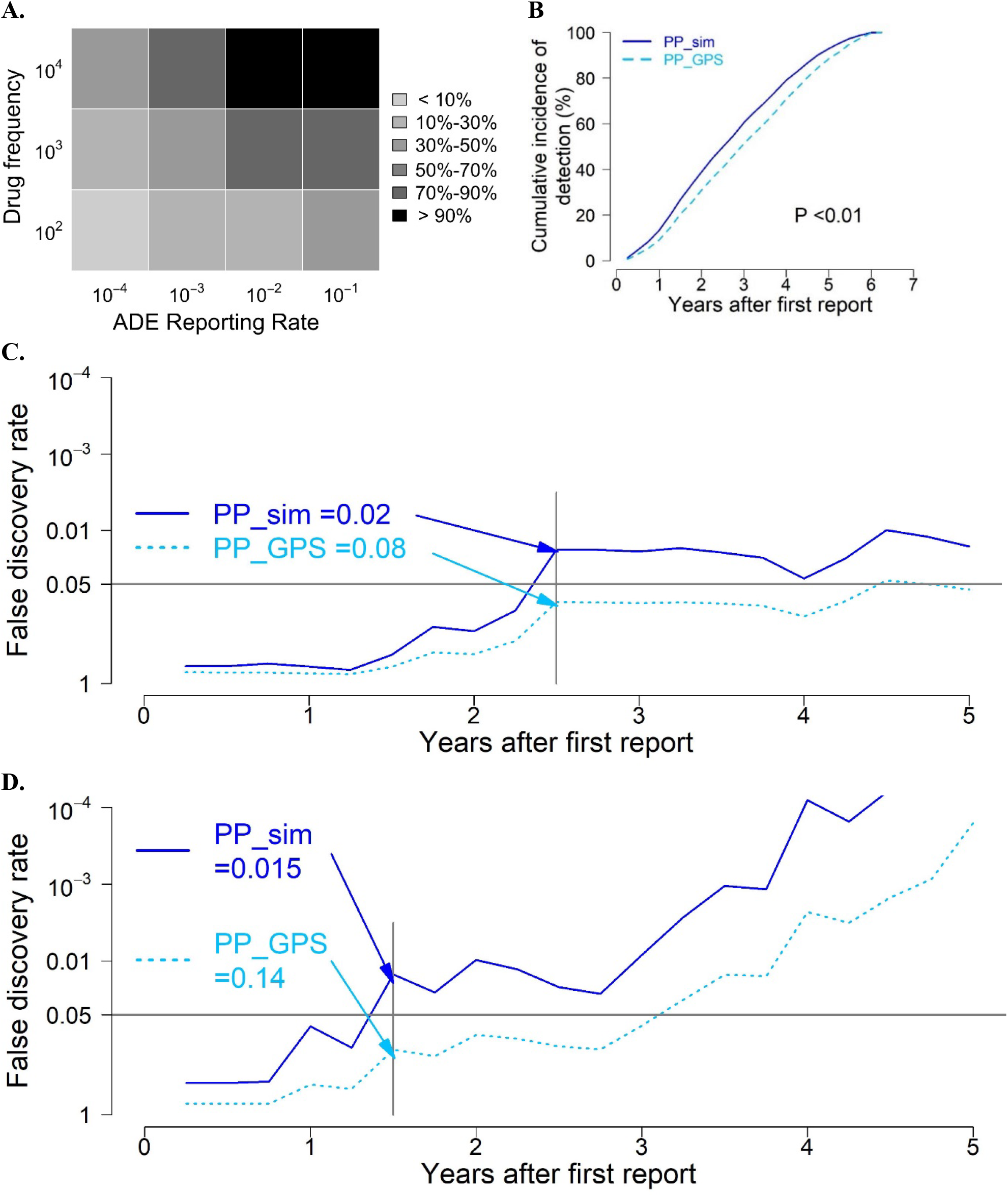
FAERS data analysis; **A**. Patterns of adverse drug events (ADE) detection stratified by reporting frequency; **B**. Cumulative incidence of ADE detection; **C**. Case study of ponatinib and hepatotoxicity; **D**. Case study of ponatinib and hepatotoxicity.

We used two examples to illustrate the use of PP_sim to sequentially monitor an ADE of a “new drug” (Figure 3C and 3D). First, Figure 3C shows PP_sim and PP_GPS for ponatinib and hepatotoxicity after ponatinib was firstly reported. Please noting that hepatotoxicity is labeled as black box warning for ponatinib.^35^ In this example, PP_sim reached the 0.05 threshold on the 10^th^ quarter after the drug’s first reporting quarter, while PP_GPS reached the 0.05 threshold on the 18^th^ quarter. In another word, PP_sim was able to identify the signal of ponatinib induced hepatotoxicity 2 years prior to PP_GPS. Second, Figure 3D shows PP_sim and PP_GPS for cabozantinib and haemoptysis after cabozantinib was firstly reported. Please noting that haemoptysis is also labeled as black box warning for cabozantinib.^35^ In this example, PP_sim reached the 0.05 threshold on the 6^th^ quarter after the drug’s first reporting quarter, while PP_GPS reached the 0.05 threshold on the 13^th^ quarter. In another word, PP_sim was able to identify the signal of cabozantinib induced haemoptysis almost 2 years prior to PP_GPS.

## 4 Conclusion and Discussion

In this manuscript, we introduce the use of drug similarity based posterior probability of null hypothesis for the early detection of adverse drug event (ADE) signals. The proposed approach has better performance metrics than existing approaches in drug-label based performance evaluation analysis, especially in the first few years after a drug has been initially reported. Additionally, the proposed approach is able to identify labeled ADEs in a timelier fashion. In simulation study, the proposed approach is able to properly control FDR, as well as has better true positive rate (TPR) and excellent empirical true negative rate (TNR). Last, we illustrate the application of the proposed approach by analyzing “new drugs” that were firstly reported on or after 2012Q1 in the FAERS data. In the exemplified analysis, the proposed approach is able to identify new ADE signals with a lower drug reporting frequency and/or a smaller ADE reporting rate compared with existing approach. We also highlights two examples that the proposed approach is able to identify boxed warnings almost 2 years prior to existing approach.

Our performance evaluation analysis shows that the proposed approach has better AUC values in the first few years after a new drug has been reported comparing to existing approaches (Figure 2A and 2B). In another word, the proposed approach outperforms existing approaches at a lower drug reporting frequency. As the time increases, the advantage of the proposed approach over existing approaches decreases (Figure 2A and 2B). In another word, as the drug report frequency increases, all approaches have similar performance. The proposed approach has better performance metrics at a lower drug reporting frequency, as it leverages drug similarity based prior risk distributions. In fact, labeled drug-ADE pairs have smaller prior probabilities of null hypothesis compared with unlabeled drug-ADE pairs (Figure S1). Such a fact demonstrates that drug similarity based prior risk distributions contribute relevant ADE risk information to ADE detection. Our findings agree with existing literatures.^23,24^ Compared with fiscal year based evaluation analysis in existing literatures,^23,24^ our time-to-event based evaluation analysis provides more comprehensive longitudinal relationship between time after first report and performance of ADE detection. Additionally, we explicitly demonstrated that the proposed approach is able to identify labeled ADE in a timelier fashion by investigating the cumulative incidence of detection (Figure 2C and 2D).

Our simulation study demonstrates that the proposed approach is able to properly control FDR. In fact, using 0.05 as a threshold of the proposed drug similarity based posterior probability of the null hypothesis yields a conservative FDR (Figure 3A). Additionally, the proposed approach has better true positive rates (Figure 3B) and excellent true negative rates (Figure S2). These performance metrics reassure the use of the proposed approach to control FDR for monitoring a large number of ADEs of multiple new drugs.

Our exemplified analysis of “new drugs” in FAERS demonstrated that the proposed approach is able to both identify new ADE signals and detect ADEs in a timelier fashion compared with existing approach (Figure 4A and 4B). The new ADE signals identified by the proposed approach are drug-ADE pairs with a lower drug reporting frequency and/or a smaller marginal ADE reporting frequency. This observation and results from our performance evaluation analysis and simulation study suggest that the proposed approach has better sensitivity on early ADE detection, while controlling FDR. We further show that the proposed approach is able to identify two black box warnings almost 2 years earlier than existing approach (Figure 4C and 4D). These specific cases further confirm that the proposed approach is able to detect ADE in a timelier fashion.

The scope of the current work is to provide an approach for early ADE detection, while controlling FDR. We want to end with one limitation that the proposed approach does not incorporate the strength of the drug (e.g. dosage level) which is important for ADE, as dosage data are not well captured in FAERS. This limitation can be addressed by implement the proposed approach to electronic health record (EHR) data. Currently, data in many EHR systems are almost real-time.^36,37^ Thus, to extent the proposed approach to EHR data is an important future direction, as doing so shall facilitate both real-time early ADE detection and precision ADE detection. However, doing so may require the implementation of rigorous epidemiology designs and ADE phenotyping algrothim.^38^

## Data Availability

All data produced in the present study are available upon reasonable request to the authors

## Acknowledgments

This work has been supported by the NIH grant R01GM141279.

## Notes

### Competing Interest Statement

The authors have declared no competing interest.

### Author Declarations

FDA Adverse Event Reporting System (FAERS) data which is available to the public.

## References

1 Nebeker, J. R., Barach, P. & Samore, M. H. Clarifying adverse drug events: a clinician’s guide to terminology, documentation, and reporting. Ann Intern Med 140, 795–801, doi:10.7326/0003-4819-140-10-200405180-00009 (2004).

2 Formica, D. et al. The economic burden of preventable adverse drug reactions: a systematic review of observational studies. Expert Opin Drug Saf 17, 681–695, doi:10.1080/14740338.2018.1491547 (2018).

3 Li, H., Deng, J., Yu, P. & Ren, X. Drug-Related Deaths in China: An Analysis of a Spontaneous Reporting System. Front Pharmacol 13, 771953, doi:10.3389/fphar.2022.771953 (2022).

4 Morimoto, T. et al. Incidence of adverse drug events and medication errors in Japan: the JADE study. J Gen Intern Med 26, 148–153, doi:10.1007/s11606-010-1518-3 (2011).

5 Stausberg, J. International prevalence of adverse drug events in hospitals: an analysis of routine data from England, Germany, and the USA. BMC Health Serv Res 14, 125, doi:10.1186/1472-6963-14-125 (2014).

6 Centers for Disease Control and Prevention (CDC). Adverse Drug Events in Adults, <https://www.cdc.gov/medicationsafety/adult_adversedrugevents.html> (2017).

7 Food and Drug Administration (FDA). Preventable Adverse Drug Reactions: A Focus on Drug Interactions, <https://www.fda.gov/drugs/drug-interactions-labeling/preventable-adverse-drug-reactions-focus-drug-interactions> (2018).

8 Sarkar, U., Lopez, A., Maselli, J. H. & Gonzales, R. Adverse drug events in U.S. adult ambulatory medical care. Health Serv Res 46, 1517–1533, doi:10.1111/j.1475-6773.2011.01269.x (2011).

9 Downing, N. S. et al. Postmarket Safety Events Among Novel Therapeutics Approved by the US Food and Drug Administration Between 2001 and 2010. Jama-J Am Med Assoc 317, 1854–1863, doi:10.1001/jama.2017.5150 (2017).

10 Santoro, A., Genov, G., Spooner, A., Raine, J. & Arlett, P. Promoting and Protecting Public Health: How the European Union Pharmacovigilance System Works. Drug Saf 40, 855–869, doi:10.1007/s40264-017-0572-8 (2017).

11 Food and Drug Administration (FDA). FDA Adverse Events Reporting System (FAERS) Public Dashboard, <https://fis.fda.gov/sense/app/95239e26-e0be-42d9-a960-9a5f7f1c25ee/sheet/7a47a261-d58b-4203-a8aa-6d3021737452/state/analysis> (2022).

12 Evans, S. J., Waller, P. C. & Davis, S. Use of proportional reporting ratios (PRRs) for signal generation from spontaneous adverse drug reaction reports. Pharmacoepidemiol Drug Saf 10, 483–486, doi:10.1002/pds.677 (2001).

13 van Puijenbroek, E. P. et al. A comparison of measures of disproportionality for signal detection in spontaneous reporting systems for adverse drug reactions. Pharmacoepidemiol Drug Saf 11, 3–10, doi:10.1002/pds.668 (2002).

14 Huang, L., Zalkikar, J. & Tiwari, R. C. Likelihood ratio test-based method for signal detection in drug classes using FDA’s AERS database. J Biopharm Stat 23, 178–200, doi:10.1080/10543406.2013.736810 (2013).

15 Bate, A. Bayesian confidence propagation neural network. Drug Saf 30, 623–625, doi:10.2165/00002018-200730070-00011 (2007).

16 DuMouchel, W. Bayesian Data Mining in Large Frequency Tables, with an Application to the FDA Spontaneous Reporting System. The American Statistician 53, doi:https://doi.org/10.2307/2686093 (1999).

17 Harpaz, R. et al. Novel data-mining methodologies for adverse drug event discovery and analysis. Clin Pharmacol Ther 91, 1010–1021, doi:10.1038/clpt.2012.50 (2012).

18 Ahmed, I. et al. Bayesian pharmacovigilance signal detection methods revisited in a multiple comparison setting. Stat Med 28, 1774–1792, doi:10.1002/sim.3586 (2009).

19 Tatonetti, N. P., Ye, P. P., Daneshjou, R. & Altman, R. B. Data-driven prediction of drug effects and interactions. Sci Transl Med 4, 125ra131, doi:10.1126/scitranslmed.3003377 (2012).

20 Harpaz, R. et al. Performance of pharmacovigilance signal-detection algorithms for the FDA adverse event reporting system. Clin Pharmacol Ther 93, 539–546, doi:10.1038/clpt.2013.24 (2013).

21 Patadia, V. K. et al. Evaluating performance of electronic healthcare records and spontaneous reporting data in drug safety signal detection. Int J Clin Pharm 37, 94–104, doi:10.1007/s11096-014-0044-5 (2015).

22 Iyer, S. V., Harpaz, R., LePendu, P., Bauer-Mehren, A. & Shah, N. H. Mining clinical text for signals of adverse drug-drug interactions. J Am Med Inform Assoc 21, 353–362, doi:10.1136/amiajnl-2013-001612 (2014).

23 Ji, X. et al. Combining a Pharmacological Network Model with a Bayesian Signal Detection Algorithm to Improve the Detection of Adverse Drug Events. Front Pharmacol 12, 773135, doi:10.3389/fphar.2021.773135 (2021).

24 Liu, R. & Zhang, P. Towards early detection of adverse drug reactions: combining pre-clinical drug structures and post-market safety reports. BMC Med Inform Decis Mak 19, 279, doi:10.1186/s12911-019-0999-1 (2019).

25 Vilar, S., Tatonetti, N. P. & Hripcsak, G. 3D pharmacophoric similarity improves multi adverse drug event identification in pharmacovigilance. Sci Rep 5, 8809, doi:10.1038/srep08809 (2015).

26 Kim, S. et al. PubChem Substance and Compound databases. Nucleic Acids Res 44, D1202–1213, doi:10.1093/nar/gkv951 (2016).

27 World Health Organization (WHO) Collaborating Centre for Drug Statistics Methodology. Purpose of the ATC/DDD system, <https://www.whocc.no/atc_ddd_methodology/purpose_of_the_atc_ddd_system/> (

28 Banda, J. M. et al. A curated and standardized adverse drug event resource to accelerate drug safety research. Sci Data 3, 160026, doi:10.1038/sdata.2016.26 (2016).

29 DrugBank, <https://go.drugbank.com/> (2022).

30 National Library of Medicine. Unified Medical Language System® (UMLS®), <https://www.nlm.nih.gov/research/umls/rxnorm/index.html> (2021).

31 Lindstrom-Gommers, L. & Mullin, T. International Conference on Harmonization: Recent Reforms as a Driver of Global Regulatory Harmonization and Innovation in Medical Products. Clin Pharmacol Ther 105, 926–931, doi:10.1002/cpt.1289 (2019).

32 Side Effect Resource (SIDER 4.1), <http://sideeffects.embl.de/> (

33 Demner-Fushman, D. et al. A dataset of 200 structured product labels annotated for adverse drug reactions. Sci Data 5, 180001, doi:10.1038/sdata.2018.1 (2018).

34 Schonbrodt, F. D., Wagenmakers, E. J., Zehetleitner, M. & Perugini, M. Sequential hypothesis testing with Bayes factors: Efficiently testing mean differences. Psychol Methods 22, 322–339, doi:10.1037/met0000061 (2017).

35 Food and Drug Administration (FDA). Drug Label of Cabozantinib, <https://www.accessdata.fda.gov/drugsatfda_docs/label/2012/203756lbl.pdf> (

36 Khurshid, A. et al. Developing a real-time EHR-integrated SDoH clinical tool. AMIA Jt Summits Transl Sci Proc 2020, 308–316 (2020).

37 Symons, J. D., Ashrafian, H., Dunscombe, R. & Darzi, A. From EHR to PHR: let’s get the record straight. BMJ Open 9, e029582, doi:10.1136/bmjopen-2019-029582 (2019).

38 Chiang, C. W. et al. Random control selection for conducting high-throughput adverse drug events screening using large-scale longitudinal health data. CPT Pharmacometrics Syst Pharmacol 10, 1032–1042, doi:10.1002/psp4.12673 (2021).

