## Supplemental Figures for "An Early Adverse Drug Event Detection Approach with False Discovery Rate Control"

### Supplementary Figures

**Figure S1.** Distributions of the prior probability of null hypothesis; **A.** ADEs in Demner-Fushman et al. (2018); **B.** ADEs in SIDER.

**Figure S2.** True negative rate (TNR) in simulation study.

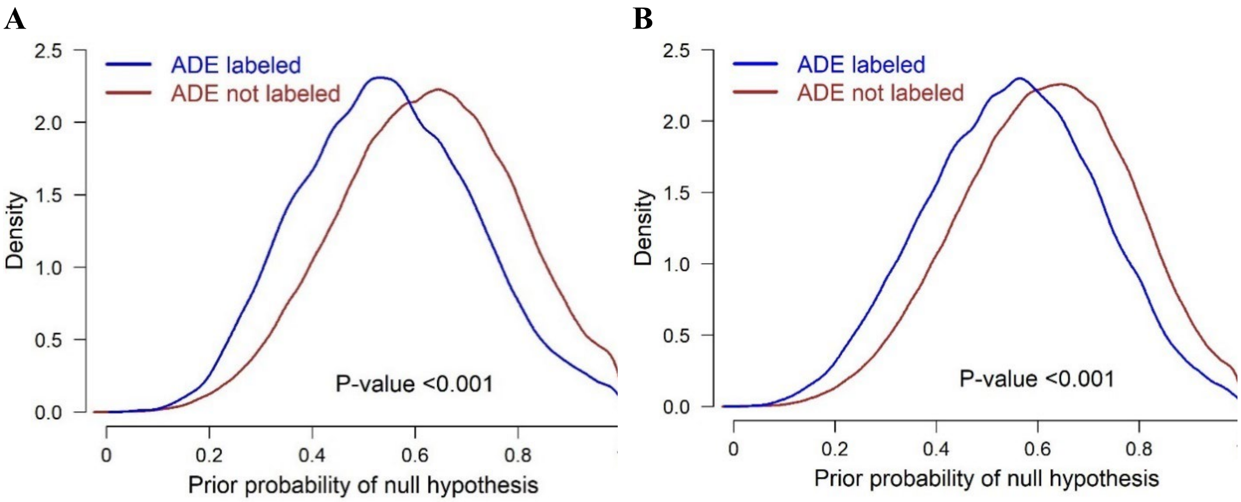


**Figure S1.** Distributions of the prior probability of null hypothesis; **A.** ADEs in Demner-Fushman et al. (2018); **B.** ADEs in SIDER.

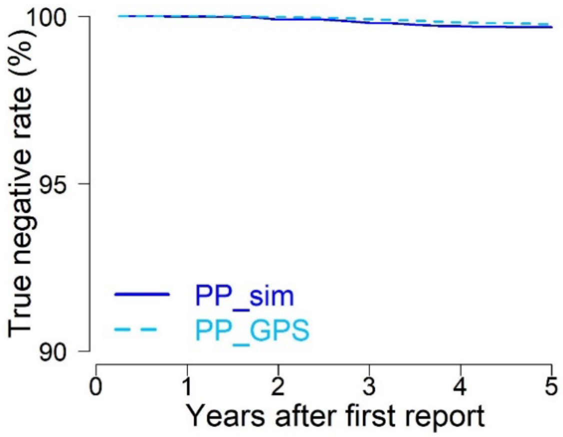


**Figure S2.** Empirical True negative rate (TNR) in simulation study.
